# When Cuts Cost More: Projected Fiscal Impact of Eliminating the AIDS Drug Assistance Program in 30 US States

**DOI:** 10.64898/2026.07.27.26359045

**Authors:** Ryan M. Forster, Melissa Schnure, Ruchita Balasubramanian, Joyce L. Jones, Emily P. Hyle, Scott Batey, Keri N Althoff, Kelly Gebo, David W. Dowdy, Maunank Shah, Anthony T. Fojo, Parastu Kasaie

## Abstract

**Background:** The AIDS Drug Assistance Program (ADAP) supports antiretroviral therapy for low-income people with HIV. We assessed whether ADAP savings would offset additional HIV care costs resulting from new infections.

**Methods:** Using the Joint HIV Epidemiologic and Economic Model calibrated to 30 US states and the District of Columbia, we compared continued 2025 ADAP coverage with elimination beginning Jan-2026. We projected excess HIV infections and care costs through 2035 under three cost-scenarios. Projected costs were limited to HIV care costs attributable to excess incident infections that would results from eliminating ADAP. The primary outcome was the Net Cost to Expenditure Ratio (NCER), defined as healthcare-system costs minus ADAP expenditure, relative to ADAP expenditure by 2035.

**Results:** Across the modeled US jurisdictions, ADAP elimination was projected to result in a median of 143,347 additional HIV incident cases, generating $11.24 billion in HIV care costs through 2035. Compared with $6.58 billion in ADAP spending, this resulted in $4.66 billion in net additional costs (NCER=0.71). For every $1 in ADAP spending avoided, the healthcare system incurred $1.71 in downstream HIV care costs, a net increase of $0.71. This pattern was strongest in states with high baseline ADAP coverage and more rural HIV populations, many of which were Medicaid non-expansion states in the South, with substantial variation in NCER across jurisdictions (3.79 in Missouri to −0.97 in the District of Columbia).

**Conclusions:** Eliminating ADAP would be medically and fiscally counterproductive, increasing HIV burden and generating healthcare costs that would exceed program savings within seven years.

## Introduction

The AIDS Drug Assistance Program (ADAP), funded through Ryan White Part B, provides access to antiretroviral therapy (ART) and insurance support for low-income people with HIV (PWH) who otherwise lack adequate coverage to receive HIV treatment. Since 2015, federal ADAP funding has remained flat in nominal dollars, while the client population grew by 56% from 2007 to 2024 and drug and insurance costs continued to rise – eroding the program’s real purchasing power [1]. Compounded by recent increases in Affordable Care Act (ACA) marketplace premium costs, at least 23 states (including the District of Columbia) have implemented or are considering cost containment strategies, including targeting ADAP eligibility, formularies, premium assistance, and enrollment caps or waitlists. Among these, Florida’s proposed restrictions have been the most substantial [2,3].

These local policy disruptions can have important epidemiological and health consequences: prior simulation studies have demonstrated that disruptions to Ryan White services can substantially undermine viral suppression and increase HIV incidence at the population level [4]. Given the lifelong treatment requirement following HIV acquisition and the substantial per-person cost of antiretroviral drugs, we hypothesized that downstream increases in healthcare utilization costs due to additional incident HIV infections could offset fiscal savings through ADAP disruption.

To test this hypothesis, we applied a previously published simulation of the HIV epidemic across 30 US states and the District of Columbia to conduct a cost-offset analysis [5]. Specifically, we compared the 10-year savings from complete ADAP elimination beginning in 2026 with projected cumulative HIV care costs attributable to excess incident infections through 2035.

## Methods

We used the Joint HIV Epidemiologic and Economic Model (JHEEM), a dynamic compartmental model of people living with HIV in the US **(Figure S1)** [6,7]. JHEEM has been calibrated to state-level HIV epidemics across 30 US states and the District of Columbia – representing over 97% of people living with HIV in the US [7] – capturing annual trends in several epidemiological indicators (such as HIV diagnosis and viral suppression), as well as key Ryan White program indicators at the state level. These indicators include the number of clients, viral suppression among outpatient ambulatory health service recipients, ratio of ADAP to non-ADAP clients, proportion of ADAP clients suppressed, all according to demographics and risk factor strata [4].

### Experimental Scenarios

We simulated two scenarios for 2026-2035: (1) continued ADAP coverage and spending at 2025 levels, and (2) ADAP elimination beginning Jan 1^st^, 2026. To estimate the impact of ADAP elimination on viral suppression, we drew on a survey of 180 Ryan White clinic directors, administrators, and public health officials, who estimated a mean decline in viral suppression of 61% among ADAP recipients in Medicaid expansion states (interquartile range [IQR]: 30-90%) and 73% (IQR: 71-100%) in non-expansion states, following ADAP elimination [4]. We fitted a multivariate kernel density distribution to these estimates and randomly sampled from the resulting distributions to incorporate uncertainty in ADAP elimination scenario.

### Costing Scope

Following ADAP elimination, four patient groups may incur costs or contribute to downstream transmission **(Figure 1, Table S1)**. ADAP recipients will fall into two categories: those who retain treatment but lose access to discounted drug pricing (group 1); and those who experience treatment interruption and viremia, increasing transmission risk (group 2). Among those newly infected, we distinguish two additional groups: those who are diagnosed and initiate ART during the projection period (group 3), and those who remain undiagnosed and intreated (group 4). We focus cost tracking on group 3, as their care costs are directly attributable to excess HIV infections resulting from ADAP elimination. Consequently, our estimates are conservative. We did not include costs associated with viral rebound, opportunistic infections, hospitalizations, untreated HIV, or higher medication prices among existing recipients who retained treatment after losing ADAP support.

**Figure 1.**
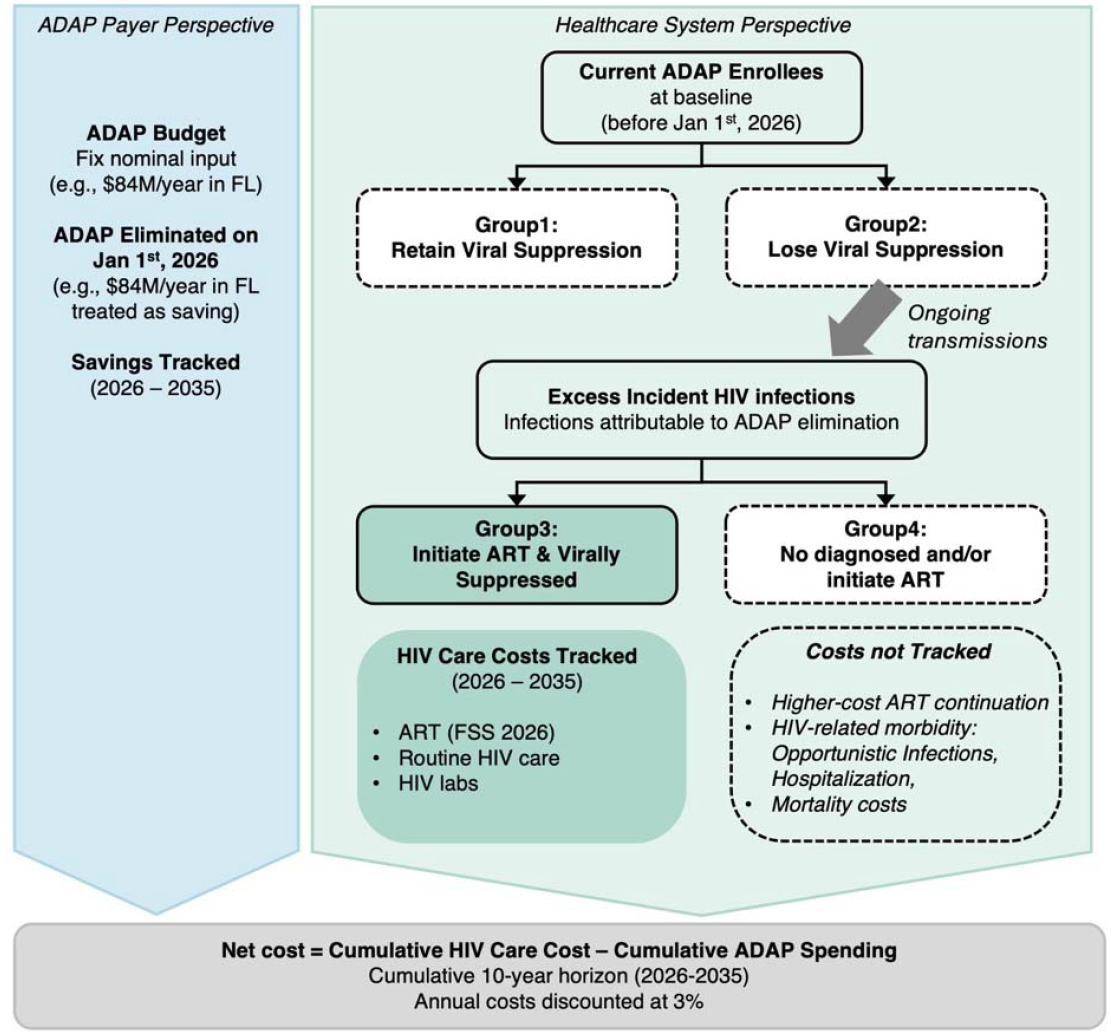
Modeling framework for ADAP elimination across 30 US States and DC, 2026–2035. ADAP elimination is assumed to occur on January 1, 2026. The blue box represents ADAP budget savings under the elimination scenario (e.g., $748.5 million/year in the US), reflecting a government-payer perspective. The green box represents epidemiological outcomes and HIV care costs from a healthcare-system perspective. Existing ADAP enrollees are stratified by viral suppression status (Groups 1 and 2), while incident HIV infections are classified as those who are diagnosed and initiate ART (Group 3) and those who remain undiagnosed and/or do not initiate ART (Group 4). HIV care costs are tracked only for Group 3 and include costs for antiretroviral therapy based on 2026 Federal Supply Schedule prices, routine clinical care, and laboratory monitoring. Dashed boxes indicate costs that are not included in the analysis; incorporating these costs would increase the estimated economic burden and reflect a broader societal perspective. Net cost is estimated as cumulative HIV care costs minus cumulative ADAP spendings over 2026–2035. Costs associated with viral rebound, opportunistic infections, hospitalizations, untreated HIV, or higher medication prices among existing recipients who retained treatment after losing ADAP support are not included.

### Costing Assumptions

Individuals engaged in HIV care and receiving ART incur costs associated with routine clinical care, laboratory monitoring, and ART medications. We estimated annual routine care costs by combining CD4-stratum utilization from Neilan et al. (2020) with 2023-unit costs from Jones et al. (2025) **(Table S2)** [3,4]. Because JHEEM does not model CD4 status, costs were population-weighted using the observed CD4 distribution among people receiving HIV care [4]. Laboratory monitoring costs assumed 2.4 CD4 and 2.4 HIV viral load tests per person-year, at $50 and $90 per unit, respectively in 2023 USD [4,5].

#### Cost of ART

We estimated ART costs for four major regimen classes, including integrase strand transfer inhibitor (INSTI)-based, protease inhibitor (PI)–based and non-nucleoside reverse transcriptase inhibitor (NNRTI)–based, and multicore regimens. We adopted the regimen classifications by Ma et al. (2022) and extracted drug costs from the 2026 Federal Supply Schedule (FSS) [6,7]. For each class, FSS prices defined a range of annual treatment costs based lowest and highest observed prices across regimens. We then combined these estimates to obtain an overall cost range for each regimen class **(Table S3)**. We constructed three ART cost scenarios using the lower bound, median, and upper bound of each range, weighted by regimen use to estimate average annual ART cost per treated individual [9].

Subsequently, total annual HIV care cost for newly incident HIV entering care and initiating ART during the projection period were estimated under three cost tiers: low=$20,900, median=$35,000, and high=$45,000 (Table S4).

All costs were adjusted to 2026 USD pricing using the Medical Care Consumer Price Index (mCPI)[5]. ART drug costs were inflated at 5.4% per year and routine care costs at 5.6% per year [8]. Baseline ADAP spending were extracted from state reports and held constant in nominal terms through 2035 [2,14]. All projected costs and ADAP spending were discounted to the start of 2026 at 3% per year as per standard health economics convention [11].

### Outcomes

The primary outcome was the 2035 **Net Cost of ADAP elimination to ADAP Expenditure ratio (NCER): NCER** 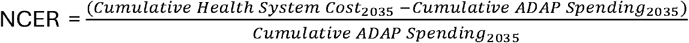 . NCER values greater than zero indicate that downstream HIV care costs exceed savings from ADAP elimination.

Both policy scenarios were simulated across 1,000 calibrated simulations under each of the three cost scenarios, and outcomes were reported as medians and 95% uncertainty ranges. All analyses were conducted in R.

### Secondary Analyses

We examined how NCER ratio varied by each state’s Medicaid expansion status; and assessed associations between projected outcomes and key state-level characteristics, including average HIV transmission rate in 2025; ADAP spending per client in 2025; proportion of people with suppressed HIV that are covered by ADAP in 2025; and urbanicity, defined as the mean proportion of residents living in urban areas across counties in 2021, weighted by each county’s share of statewide diagnosed HIV prevalence. Spearman’s rank correlations were used to assess associations.

Separately, we carried out a threshold analysis to identify when eliminating ADAP would shift from cost-saving to cost-incurring by 2035. To do this, we changed two factors separately: the proportion of ADAP clients who lost viral suppression after ADAP elimination, and annual per-person ART costs. For viral suppression, we reran the simulations assuming 10%–100% of ADAP-supported, virally suppressed clients lost viral suppression following ADAP elimination in 2026, in 10-percentage-point increments. For ART costs, we varied 2026 annual per-person costs from

$1,000 to $100,000 on a logarithmic grid. In each scenario, we calculated the proportion of simulations in which downstream HIV costs exceeded eliminated ADAP spending (NCER > 0) and identified the threshold at which more than 50% of simulations became cost-incurring **(Supplement-S1.5)**.

## Results

### National trends

Across all 30 modeled states and the District of Columbia, ADAP elimination was projected to result in a median of 112,457 [95% Uncertainty Range (UR): 12,590–178,708] individuals losing viral suppression, leading to 143,347 [16,946–226,886] additional incident HIV infections between Jan 2026 and Dec 2035 **(Table 1)**. These individuals were projected to accumulate $11.24B [$1.67B–$22.66B] in downstream HIV care costs by 2035, compared against $6.58B saving from ADAP elimination **(Figure 2A)**. Only $27.7M [$3.8M–$57.9M] of these costs were realized in the first year, compared to $748.5M expected spending from the state’s ADAP budget **(Table S6)**. However, the projected health system costs grew faster than savings, surpassing them by 2033, and reaching a net cost-to-expenditure ratio (NCER) of 0.71 [−0.75–2.45] by 2035 (i.e., for every $1 saved from ADAP elimination, the healthcare system incurred $1.71 in downstream HIV care costs, a net increase of $0.71 - **Figure 2B)**.

**Table 1.**
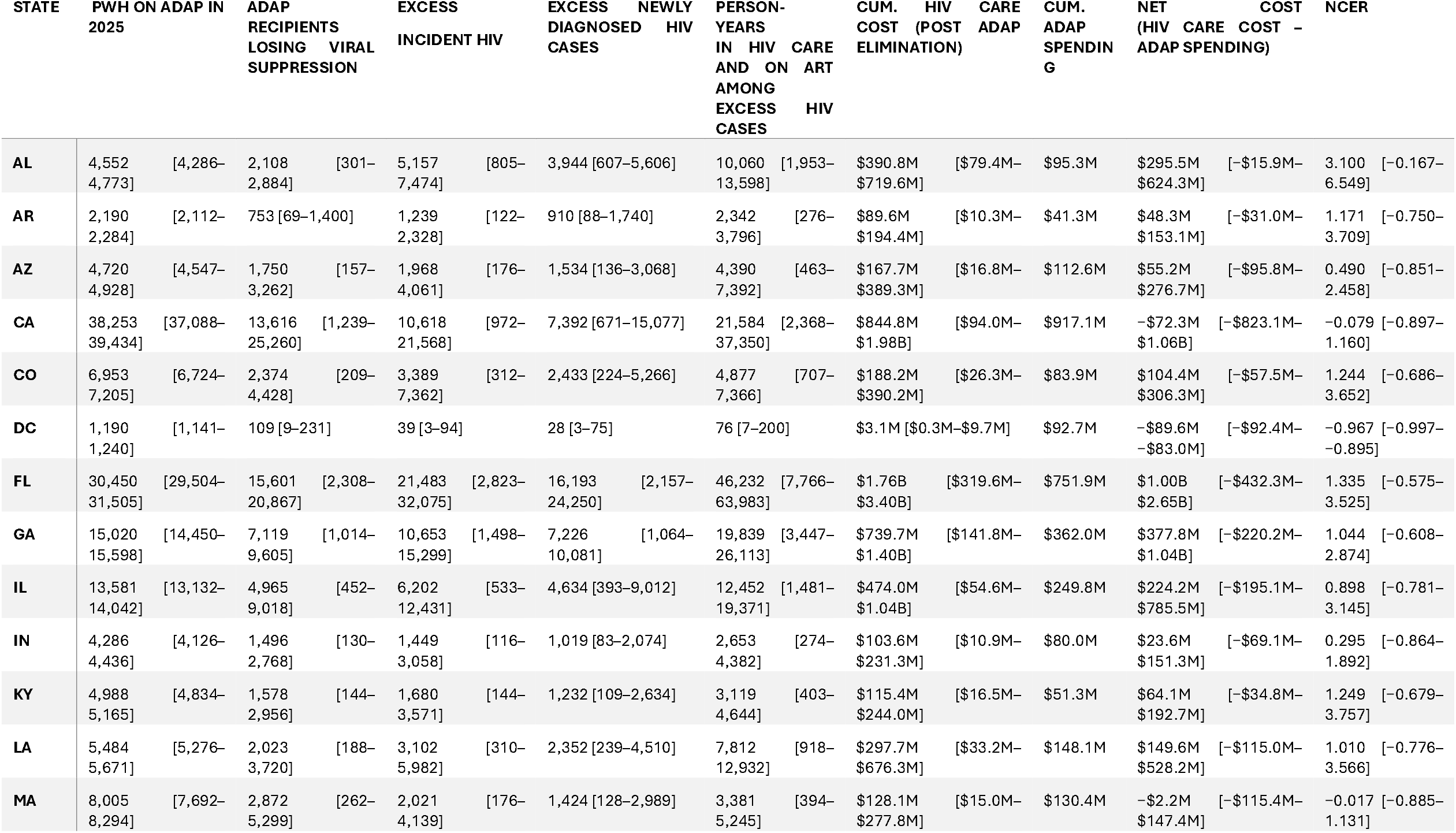
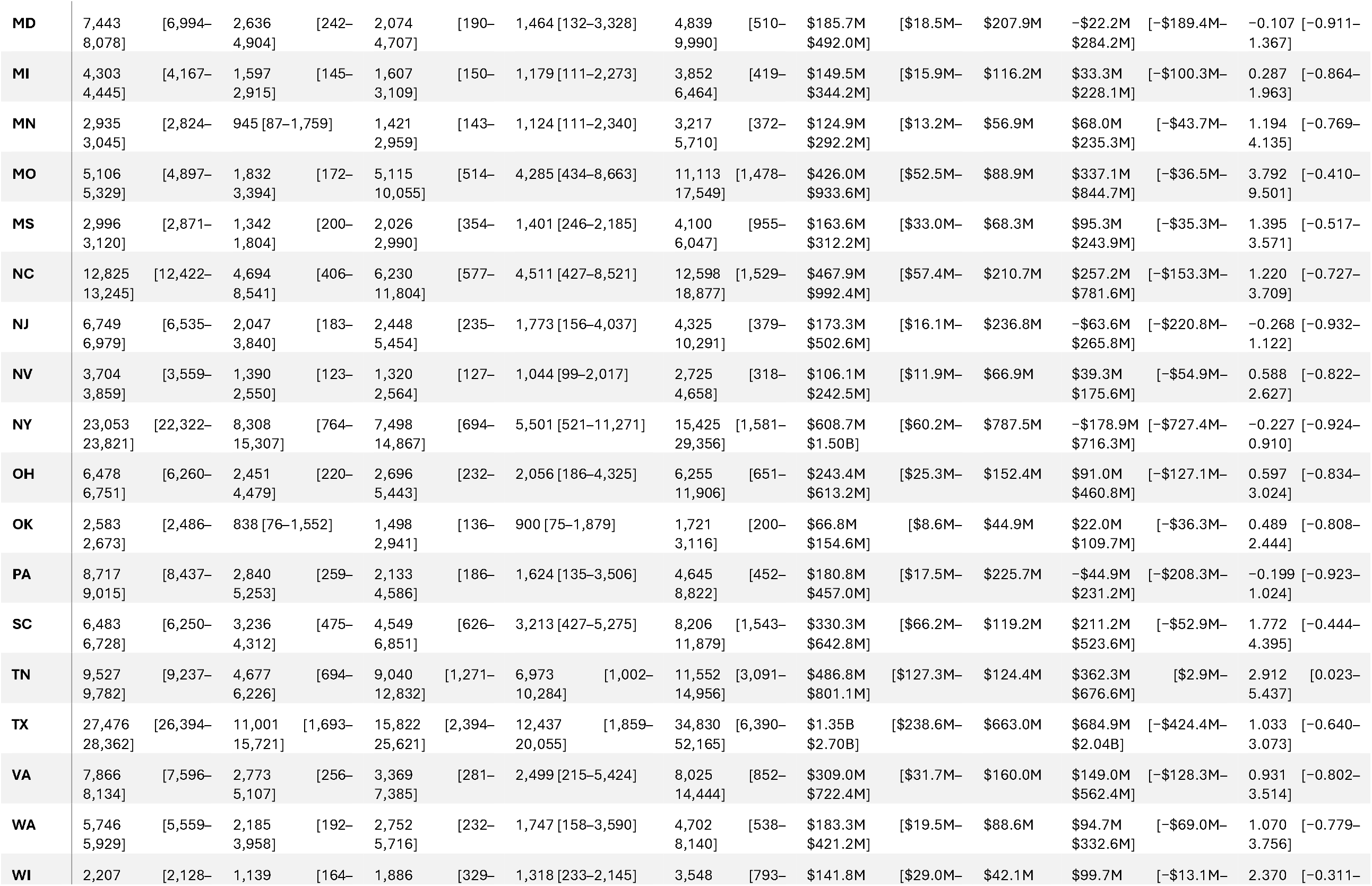
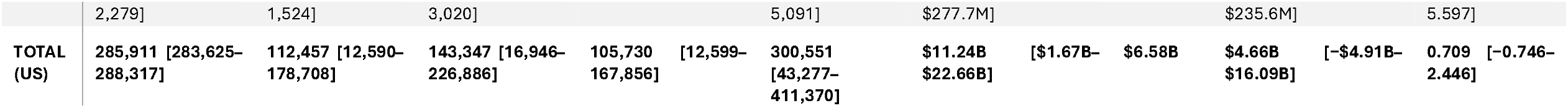
State-Level Impact of ADAP Elimination, Cumulative Through 2035.

**Figure 2.**
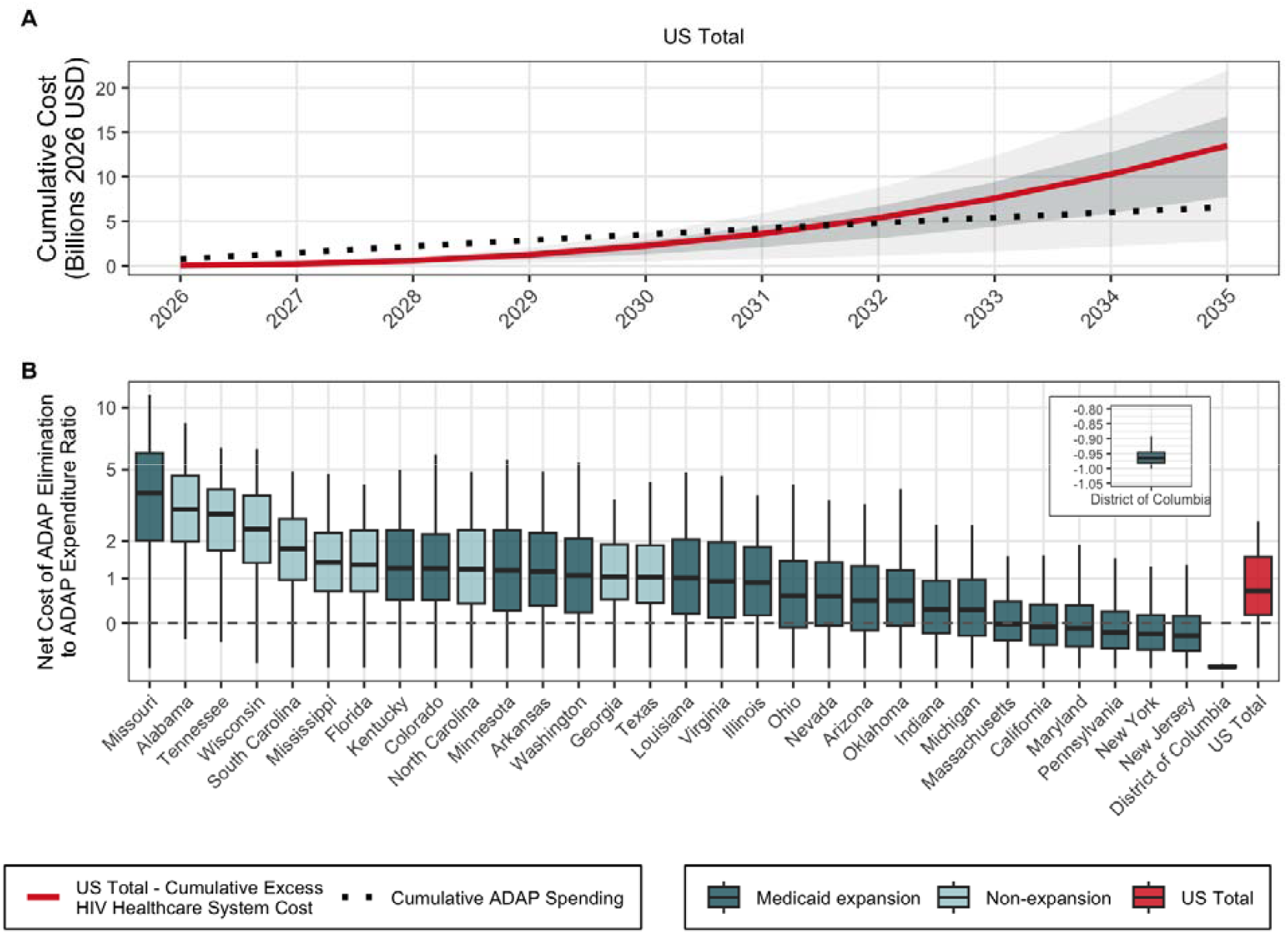
Projected HIV care cost attributable to excess new infections following elimination of the AIDS Drug Assistance Program (ADAP), 2026–2035. **(A)** Projected cumulative cost of HIV care (health system perspective) attributable to excess HIV infections entering HIV care and receiving ART across the 30 states and the District of Columbia following ADAP elimination (solid line, median cost scenario) compared to cumulative Ryan White Part B ADAP spending (dotted line) from 2026-2035. Dark and light-shaded bands indicate the 50% and 95% uncertainty intervals across transmission and cost-gradient simulations, respectively. (B) Net Cost of ADAP Elimination relative to ADAP spending ratio (NCER) across all modeled jurisdictions by 2035 (cumulative cost minus cumulative ADAP spending, divided by cumulative ADAP spending). States are ordered by median ratio (high to low) and shaded by Medicaid expansion status. The dashed horizontal line indicates cost-neutrality (ratio = 0); values above zero indicate that projected HIV care costs exceed savings from ADAP elimination. All costs are discounted at 3% annually to 2026 USD.

### Projected Epidemiologic Impact

Projected epidemiological consequences of ADAP elimination at the national level were disproportionately driven by a small number of jurisdictions: Florida was projected to experience the largest loss in viral suppression, with a projected median of 15,601 individuals becoming virally unsuppressed, followed by California (13,616), Texas (11,001), New York (8,308) and Georgia (7,119) – **Table1**. These five jurisdictions accounted for nearly half (49.5%) of the national total loss in viral suppression. Florida was also projected to experience the largest absolute increase in new HIV infections, with a projected median of 21,483 excess infections by 2035, followed by Texas (15,822), Georgia (10,653) and California (10,618). At a national level, elimination of ADAP reduced viral suppression from a projected median of 80% to 65% by 2035, nearly doubling the number of people with HIV who were not virally suppressed, from a median of 225,000 to 425,000 **(Table S5)**.

### Projected Net Costs

Before accounting for eliminated ADAP spending (savings), projected downstream HIV care costs attributable to excess infections were highest in Florida, with a median of $1.76 billion over 10 years, followed by Texas ($1.35 billion), California ($844.8 million), Georgia ($739.7 million), and New York ($608.7 million). Collectively, these five states accounted for 53% of total savings from ADAP elimination ($3.48 billion of $6.58 billion) and 47% of downstream HIV care costs ($5.30 billion of $11.24 billion)-**Table 1**. After eliminated ADAP spending was subtracted from costs, Florida had the highest projected net cost at $1.00 billion, followed by Texas ($685 million), Georgia ($378 million), Tennessee ($362 million), and Missouri ($337 million). California and New York had negative net costs (−$72.3 million and −$179 million, respectively), as did four other states and the District of Columbia, indicating that savings from ADAP elimination exceeded the downstream HIV care costs across these seven states through 2035 (**Table 1**).

### Projected Net Cost to Expenditure Ratio

Expressing these projected net costs relative to ADAP savings (via NCER) provides a size-independent basis for cross-jurisdictional comparison. Missouri had the largest projected median NCER at 3.79, followed by Alabama (3.10), Tennessee (2.91) and Wisconsin (2.37) – indicating that in these jurisdictions, each dollar of ADAP spending avoided was associated with $2.4-$3.8 in additional downstream HIV care costs (even after accounting for ADAP savings). The District of Columbia had a projected NCER of -0.97, suggesting minimal downstream costs due to ADAP elimination (i.e., NCER of -1 implies no downstream costs). Six additional jurisdictions had negative NCER values, ranging from -0.27 to -0.02, indicating that projected savings from eliminating ADAP exceeded projected downstream costs due to excess infections from 2026-2030. Uncertainty was wide throughout: Tennessee was the only jurisdiction whose 95% uncertainty interval excluded zero (95%UR: 0.023–5.437), indicating that fewer than 2.5% of simulations showed ADAP elimination as cost-saving.

### Factors Associated with Net Cost to Expenditure Ratio

NCER estimates correlated with underlying structural differences between jurisdictions. Southern states were characterized by a higher proportion of suppressed PWH receiving ADAP support (range 26.3–61.3%), lower ADAP spending per client ($1,170–$3,074), and the highest projected NCERs (0.489–3.100) (**Figure 3**). In contrast, Northeastern states had a lower proportion of virally suppressed PWH receiving ADAP support (23.7–47.4%), greater ADAP spending per client ($1,855–$3,992), and generally lower NCERs, ranging from −0.27 in New Jersey to −0.02 in Massachusetts (**Figure 3**). The District of Columbia ranked lowest among the 31 modeled jurisdictions in the proportion of virally suppressed PWH receiving ADAP support (3.3%) and highest in ADAP spending per client ($8,866 in 2026 USD) and had the lowest NCER, as above. Massachusetts represented a partial exception to this pattern: despite relatively low ADAP spending per client ($1,855) and comparatively high reliance on ADAP among virally suppressed PWH (47.4%), it had a slightly negative median NCER (−0.05). Jurisdictions in which a smaller proportion of diagnosed PWH reside in urban counties had higher projected NCERs, ranging from an NCER of 1.395 in Mississippi (54.3% urban) to an NCER of −0.967 in the District of Columbia (100% urban). Non-Medicaid expansion states had a higher projected NCER (median 1.395) than Medicaid expansion states (median 0.539) (**Table S7**). State-level variation in NCER was weakly correlated with the underlying transmission rate, viral suppression, and diagnosed HIV prevalence, although there was a moderate correlation with cumulative ADAP spending (**Figure 3 & S2**).

**Figure 3.**
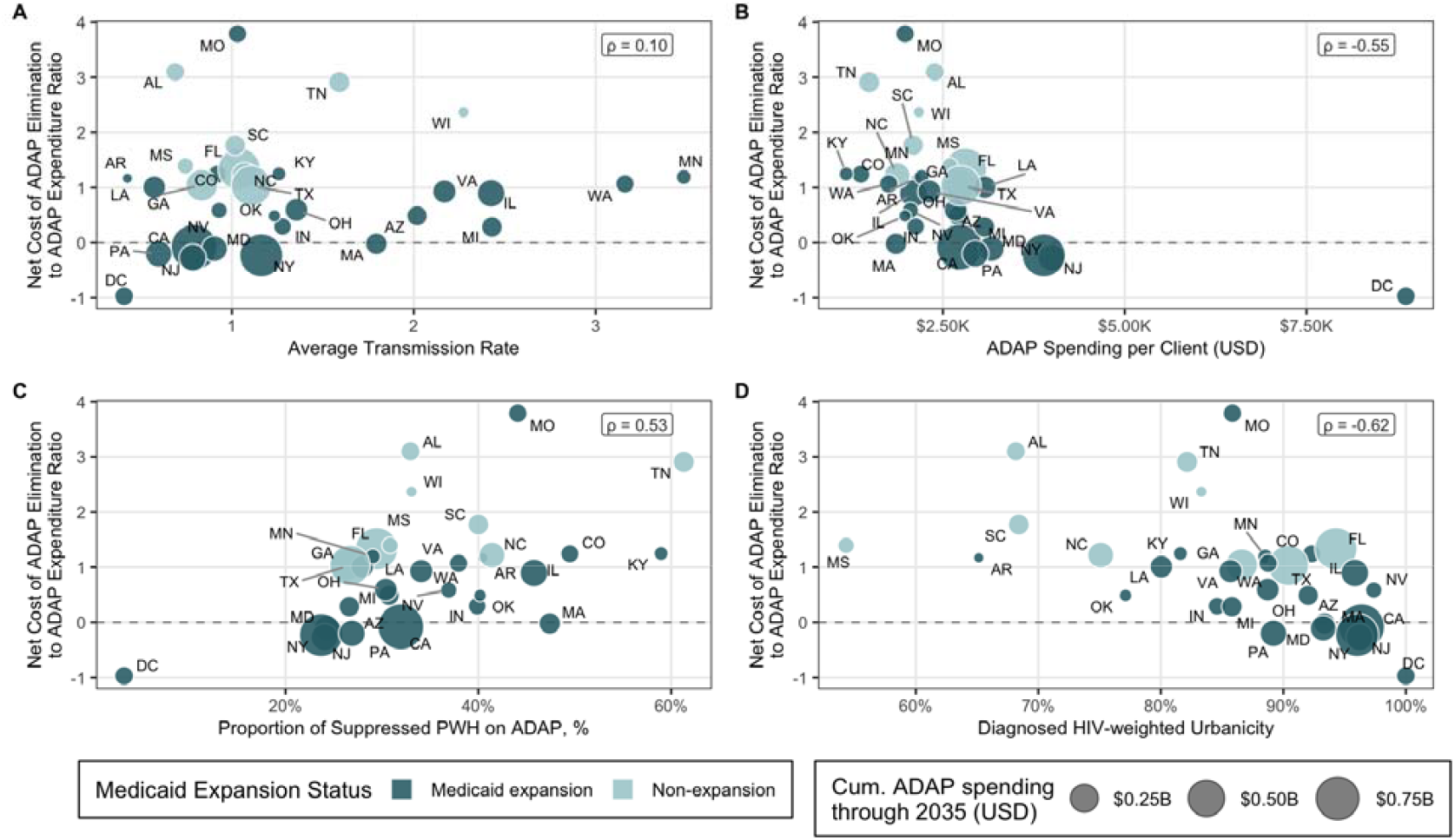
Association between the net cost of ADAP elimination (relative to ADAP spending) ratio and selected state-level characteristics. The Y-axis represents the projected net cost of ADAP elimination relative to baseline ADAP spending by 2035. Each panel shows the correlation with a different state-level characteristic: (A) average HIV transmission rate in 2025; (B) ADAP spending per client in 2025; (C) proportion of people with suppressed HIV that are covered via ADAP in 2025; and (D) urbanicity, defined as the mean proportion of county residents living in urban areas weighted by each county’s share of statewide diagnosed HIV prevalence in 2021. Point size I proportional to cumulative ADAP spending through 2035, if ADAP spending were not eliminated, and point color indicates Medicaid expansion status. The symbol ρ denotes the Spearman correlation coefficient. The dashed horizontal line indicates cost-neutrality (ratio = 0); values above zero indicate that projected care costs exceed ADAP spending.

### Threshold Analysis

In the primary analysis, ADAP elimination was projected to be cost-incurring (i.e., NCER>0) in more than 50% of simulations in 24 of 31 modeled jurisdictions; the District of Columbia was the only jurisdiction that remained cost-saving across all simulations (**Figure S3A**). Non-Medicaid expansion states were more likely to experience higher costs if more people lost viral suppression. In most of these states, costs exceeded savings when over 13%–29% of people lost viral suppression or when annual ART costs exceeded $4,600–$12,800 (**Figures S3B&C**). Results varied more widely among Medicaid expansion states, with the levels at which costs exceeded savings ranging from 17% to 71% for loss of viral suppression and from $3,300 to $43,500 for annual ART costs. Tennessee and Missouri had higher costs than savings across all levels of viral suppression loss we examined, while the District of Columbia had savings that exceeded costs throughout.

## Discussion

Across the 30 modeled US states and the District of Columbia, for every $1 in ADAP spending eliminated beginning January 1, 2026, the healthcare system was projected to incur $1.71 in downstream HIV care costs attributable to excess incident infections by December 31, 2035, yielding a net cost increase of $0.71. This pattern was most pronounced in states with high baseline ADAP coverage and low ADAP spending per client. These states, many of them Medicaid non-expansion states in the South, also bear a disproportionate share of HIV burden in the US. These findings suggest that the short-term fiscal savings of ADAP elimination are transient, with downstream HIV care costs exceeding program savings within 5 to 9 years across all modeled states (median: 7 years), except for the District of Columbia and 6 US states.

These findings build directly on prior work demonstrating that Ryan White disruptions increase HIV incidence by adding an explicit fiscal dimension: ADAP cuts do not merely harm patients; they are fiscally counterproductive [4,7]. ART-mediated viral suppression prevents onward transmission, and ADAP is the financing mechanism that makes population-level suppression achievable among low-income PWH [16,17]. Previous studies have shown ADAP’s significant contribution to viral suppression: From 2015–2022, ADAP clients accounted for nearly one-third of all virally suppressed people in the US, despite representing less than one-quarter of PWH, indicating substantially higher viral suppression rates among ADAP clients [18]. ADAP’s effectiveness in achieving viral suppression is even more significant, despite the program’s relatively flat funding over the past decade. When that financing is removed, viral suppression declines, and transmission events accrue, each generating lifetime treatment costs that diminish the short-term fiscal savings.

States with more rural HIV populations were projected to face higher care costs relative to ADAP savings (ρ = −0.62), consistent with Balasubramanian et al. (2026), who found that states with more rural HIV epidemics faced larger incidence increases following disruptions to CDC-funded HIV testing [19]. Although our model did not explicitly represent urban-rural differences in HIV program or care delivery, these differences were indirectly captured through state-level demographics and epidemiological targets. States where diagnosed HIV is more geographically concentrated in urban areas appear better positioned to absorb ADAP disruptions, likely reflecting stronger urban care infrastructure and greater provider density [20]. More rural states also disproportionately overlap with non-Medicaid expansion states, where the safety net is thinner, and HIV burden is concentrated among populations with limited access to alternative players. Notably, the provider survey informing our scenarios suggested a larger decline in viral suppression in non-Medicaid expansion states following ADAP elimination. Projected NCERs were also higher in states where a larger share of the virally suppressed population received ADAP services (ρ = 0.53). Together, these patterns suggest that states with lower baseline access to HIV care – characterized by greater rurality, non-expansion of Medicaid, and greater reliance on ADAP among the virally suppressed – face disproportionately worse net cost-to-expenditure ratios following ADAP elimination.

The net-cost-to-expenditure ratio (NCER) follows the same ratio structure as the return-on-investment (ROI) metric standard in HIV program economic evaluation [21,22], but expresses net cost (rather than savings) relative to the program’s spending. This provides a standard metric for comparing the fiscal consequences of eliminating ADAP across US states with varying program sizes and epidemic profiles. However, it should not be interpreted in isolation from epidemiological outcomes. For example, ADAP elimination in California is projected to result in over 10,000 excess HIV infections and $844.8 M in downstream ART care costs by 2035, yet given California’s large ADAP budget ($917.1 M in cumulative spending over the projection period), savings from ADAP elimination approximated costs attributable to downstream infections over a 10-year time horizon. This near-neutral fiscal result, however, should not be equated with epidemiological neutrality: elimination of ADAP was projected to result in over 10,000 excess HIV infections, nearly doubling the number of PWH having unsuppressed viral loads by 2035. A complete assessment of the impact of eliminating ADAP should also incorporate these health-related consequences.

We modeled ADAP elimination as an extreme scenario, whereas proposed policies generally involve partial funding reductions. If reductions in ADAP spending and their epidemiologic consequences scale proportionally, the NCER would be expected to remain relatively constant even though the absolute savings, infections, and downstream costs would vary with the magnitude of the cut. Our analyses suggested that, in most states, ADAP elimination would incur net costs within 10 years if the proportion of ADAP-supported, virally suppressed recipients who lost viral suppression exceeded a state-specific threshold. Where we identified a crossover, these thresholds ranged from 13% to 29% in Medicaid non-expansion jurisdictions and from 17% to 71% in expansion jurisdictions. Thus, even partial disruptions to viral suppression could be fiscally counterproductive. These thresholds represent conditional break-even points rather than direct projections of any specific partial-cut policy.

Several features of our analysis likely cause our projections to underestimate true downstream costs. First, we did not include costs attributable to current ADAP enrollees who may lose program access, including costs associated with viral rebound, opportunistic infections, hospitalizations, or comorbidity management among those discontinuing ART. We also did not model cost changes among enrollees who remain on ART but lose access to discounted drug pricing through ADAP, such as 340B and other subsidy programs, and would therefore obtain their existing regimens at higher Federal Supply Schedule or wholesale acquisition cost prices. Second, the ART costs in this analysis were based on Federal Supply Schedule pricing, which is lower than the prices most non-federally insured individuals and payers face [23]. Third, we do not include end-of-life care costs for individuals whose HIV progresses because of treatment interruption, costs which can be substantial [24]. Fourth, among excess incident cases, we do not include costs incurred before initial ART initiation (e.g., opportunistic infection costs). We also conservatively assumed that individuals who did not initiate ART at diagnosis would not subsequently initiate treatment during the projection period; in practice, some of these delayed initiators would be expected to enter care later and generate additional ART and HIV care costs within the 10-year cost horizon. Each of these assumptions pushes our current cost projections in a conservative direction. Finally, we did not model downstream behavioral effects of care disruption, such as disengagement from testing or prevention services, which can further amplify HIV incidence and worsen the projected fiscal balance. Accordingly, negative median NCERs in some jurisdictions should not be interpreted as evidence that ADAP elimination would produce overall cost savings, given the direct and downstream costs excluded from our analysis.

With 23 states restricting ADAP amid flat federal funding, policymakers should consider not only immediate budget savings but also the downstream HIV care costs that accumulate over time. Our findings suggest that, among 30 states and the District of Columbia, the future costs of lost viral suppression because of eliminating ADAP could exceed direct savings within 7 years, resulting in a projected $4.66 billion fiscal loss from downstream healthcare costs over 10 years. These findings can help inform discussions about the value of sustaining this essential component of the HIV care continuum.

## Data Availability

All data produced in the present work are contained in the supplement and are linked in the manuscript.

https://jheem.org/ryan-white-costing

https://github.com/CIPHER-Epi/jheem_analyses

https://github.com/CIPHER-Epi/jheem2

## Data availability

Data and model resources supporting the findings of this analysis are publicly available through two complementary platforms. The GitHub repositories https://github.com/CIPHER-Epi/jheem_analyses and https://github.com/CIPHER-Epi/jheem2 provide access to the model code, input data, and simulation outputs used to generate the results presented in this study. In addition, a user-friendly web-based application (https://jheem.org/ryan-white-costing) allows users to explore projected outcomes and access state-level results through an interactive interface.

## Acknowledgement

The authors thank Nicholas Sizemore for his help with developing the web tool, Zoe Dansky for her work curating input data, and Andrew Zalesak for his contributions to model development. They thank the 180 Ryan White clinic directors, administrators, and public health officials who responded to the survey informing this analysis.

