## Supplemental Document for "When Cuts Cost More: Projected Fiscal Impact of Eliminating the AIDS Drug Assistance Program in 30 US States"

### S1. Methodological Details

#### S1.1. Model review

We used the Johns Hopkins Epidemiological and Economic Model (JHEEM, **Fig S1**), a dynamic compartmental model of HIV transmission previously described [1]. Briefly, we model the effect of ADAP elimination on HIV incidence through changes in viral suppression, where a proportion of ADAP recipients who lose access to ADAP will discontinue antiretroviral therapy and no longer be virally suppressed. This reduction in viral suppression is informed by a survey of Ryan White clinic directors and administrators [2]. People with viremia can transmit HIV, leading to new infections. Excess new infections each year are computed as the difference in projected new HIV infections between the ADAP-elimination scenario and the no-intervention arm, aggregated across all demographic strata and 1,000 simulations.

JHEEM stratifies the adult population by age, race/ethnicity, sex and sexual behavior, and injection-drug-use history, and tracks individuals from HIV acquisition through diagnosis, viral suppression, and onward transmission. The calibrated simulations reproduce state-level trends in HIV diagnoses and viral suppression and Ryan White program indicators, including client volume, viral suppression among outpatient recipients, ADAP reliance, and viral suppression among ADAP clients [1,2].

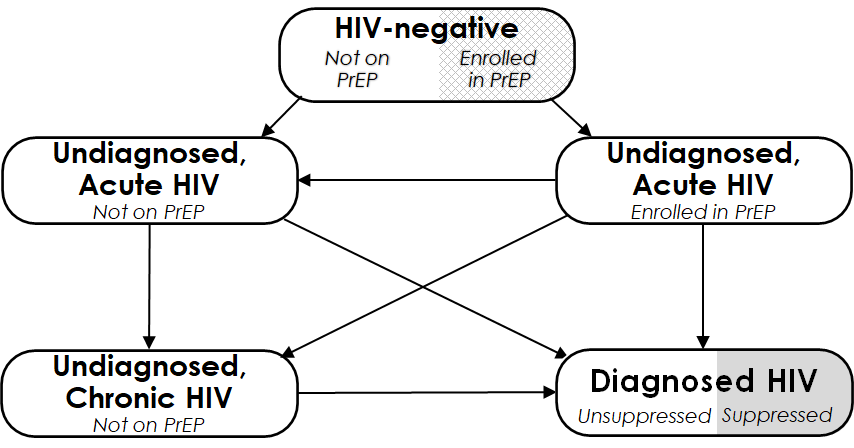

**Figure S1. JHEEM Model structure.** This figure depicts the compartments representing HIV status. Each of the five compartments is further stratified by age (13–24, 25–34, 35–44, 45–54, and ≥55 years), race/ethnicity (Black, Hispanic, and other), sex and sexual behavior (female, heterosexual male, and men who have sex with men (MSM), and intravenous drug use history (never used, active use, and prior use). “Acute HIV” refers to the first 2.9 months following infection, during which the risk of transmission is high

#### S1.2. Costing Framework

Following the elimination of ADAP funding, four patient groups may incur costs or contribute to downstream transmission (**Table S1**). We assumed that a proportion of individuals with HIV currently receiving support through ADAP retain access to care and remain virally suppressed (**Group 1**) but may incur substantially higher treatment costs if access to 340B or ADAP-negotiated drug pricing is lost and medications must instead be obtained through Federal Supply Schedule or private insurance pricing. The remaining individuals receiving support through ADAP are assumed to lose access to treatment (**Group 2**), resulting in treatment interruption, viremia, increased transmission risk, and potential health deterioration.

**Table S1. Simulated population groups impacted by ADAP elimination.**

| **Label** | **Description** | **Contribute to new infections?** | **Cost tracked?** |
| --- | --- | --- | --- |
| **Group 1: Retained in Care After ADAP Loss** | Individuals living with HIV at the time of the ADAP elimination who maintain access to care and remain on ART, potentially at higher treatment costs. | NO | NO |
| **Group 2: Lost to Care After ADAP Loss** | Individuals living with HIV at the time of the ADAP elimination who lose access to treatment, become disengaged from care, and may contribute to ongoing transmission. | YES | NO |
| **Group 3: Incident Infections Linked to HIV Care** | Newly infected individuals who are diagnosed and initiate ART during the modeling horizon | NO | YES |
| **Group 4: Incident Infections Out of Care** | Newly infected individuals who remain undiagnosed or never start ART during the modeling horizon | YES | NO |

In the ADAP elimination scenario, we assumed that the absence of ADAP support would reduce the proportion of newly diagnosed individuals who initiate ART and achieve viral suppression. This reduction was modeled by sampling a viral suppression loss parameter from its corresponding probability distribution. he distribution was estimated from survey responses among clinicians and Ryan White program directors and was specified separately for Medicaid expansion and non-expansion states, with median sampled losses of 55.7% in expansion states and 77.6% in non-expansion states.

We focus cost tracking on **Group 3**, newly infected individuals who are diagnosed and initiate ART during the projection period, because these are the people whose care costs are directly attributable to excess HIV transmission resulting from the ADAP funding cut. Finally, **Group 4** consists of newly infected individuals who remain undiagnosed or do not initiate ART during our projection period. However, they contribute to onward transmission; they do not enter the healthcare system during the modeling horizon and therefore generate no trackable HIV care costs. Consequently, our estimates are conservative, as we restrict cost calculations to the cumulative HIV care costs of excess incident cases who link to care and initiate ART during the projection period. Excess incident cases who are diagnosed but do not initiate ART in the year of diagnosis accrue no costs in this analysis, and no costs of untreated HIV disease are included for any group.

For excess incident cases who initiated ART, we assumed alternative payers (such as Medicaid or private insurance) would finance care at costs comparable to Federal Supply Schedule prices. We did not include costs associated with viral rebound, opportunistic infections, hospitalizations, untreated HIV, or higher medication prices among existing recipients who retained treatment after losing ADAP support.

#### S1.3. Cost Inputs and Assumptions

Individuals engaged in HIV care and receiving ART incur costs associated with routine clinical care, laboratory monitoring, and ART medications. We adopted estimates of routine care utilization by CD4 stratum from Neilan et al. (2020) and combined these with unit costs per visit reported by Jones et al. (2025) in 2023 USD to estimate annual routine care costs (**Table S2**) [3,4]. Because JHEEM does not explicitly model CD4 status, routine care costs were calculated as a population-weighted average across CD4 strata, using the distribution of CD4 strata observed among people receiving HIV care [4].

**Table S2. CD4 Population distribution and routine care utilization for PWH in care and receiving ART, based on Neilan et al (2020) [4].**

| **CD4 Stratum** | **% of population** | **Inpatient days per person-year** | **Outpatient Visits Per Year** | **Emergency Department Visits Per Year** |
| --- | --- | --- | --- | --- |
| **CD4 >500** | 54% | 0.3 | 6.0 | 0.3 |
| **CD4 200–500** | 37% | 0.5 | 6.0 | 0.3 |
| **CD4 <200** | 9% | 5.0 | 6.5 | 0.4 |

Laboratory monitoring costs were estimated assuming an average of 2.4 CD4 tests and 2.4 HIV viral load tests per person-year, with unit costs of $50 and $90 per test in 2023 USD, respectively [4,5]. These routine care and laboratory monitoring costs were adjusted to 2026 USD pricing using the Medical Care Consumer Price Index (mCPI)[5]. Costs were inflated according to:

$$C_{care,2026}= C_{care,2023}\times(\frac{mCPI_{2026}}{mCPI_{2023}})$$

$mCPI_{2023}=549.084$ , $mCPI_{2026}=591.677$

where $C_{\text{care},2023}$ is the reported cost in 2023 USD, $\text{mCPI}_{2023}$ and $\text{mCPI}_{2026}$ are the Medical Care Consumer Price Index values for 2023 and 2026, respectively, and $C_{\text{care},2026}$ is the inflation-adjusted cost expressed in 2026 USD.

**Cost of ART:** ART regimens have changed substantially over time, with integrase strand transfer inhibitor (INSTI)-based regimens becoming the predominant standard of care in recent years. The CFAR Network of Integrated Clinic Systems (CNICS) cohort documented high uptake of second-generation INSTI-based regimens among people with HIV in care in 2019–2020, particularly among antiretroviral-naive individuals initiating ART [6]. Accordingly, we estimated ART costs for four major regimen classes, including integrase strand transfer inhibitor (INSTI)-based, protease inhibitor (PI)–based and non-nucleoside reverse transcriptase inhibitor (NNRTI)–based, and multicore regimens. We adopted the regimen classifications by Ma et al (2022) and extracted corresponding drug costs from the 2026 Federal Supply Schedule (FSS) [6,7]. For each regimen class, we used FSS prices to define a range of annual treatment costs based on observed floor and ceiling prices across included regimens. We then combined these estimates to obtain an overall cost range for each regimen class (**Table S3**).

Finally, we constructed three ART cost scenarios. The **low ART cost scenario** used the lower bound of the estimated cost range for each regimen class, the **high ART cost scenario** used the upper bound, and the **median ART cost scenario** used the median of the range. For each scenario, costs were weighted by the distribution of ART regimen use to estimate the average annual ART cost per treated individual.

**Table S3. Monthly ART costs based on FSS pricing.** The range represents the range of prices for various regimens within each class, based on Ma et al (2022).

| **ART class** | **FSS Monthly range (2026 USD)** | **% PWH on each regimen** |
| --- | --- | --- |
| **INSTI-based regimens** | $1467 – $3749 | 74.2% |
| **PI-based regimens** | $2845 – $4693 | 6.1% |
| **NNRTI-based regimens** | $2445 – $3488 | 8.5% |
| **Multi-core regimens** | $2408 – $3423 | 11.2% |

The three cost components – routine care, laboratory monitoring, and ART medications – were combined to calculate the total HIV care cost incurred by newly incident HIV cases who enter care and initiate ART during the projection period across three cost tiers: low=$20,900, median=$35,000, and high=$45,000 per year (**Table S4**).

**Table S4. Total care costs in each cost tier (2026 USD).** All costs are rounded to the nearest 100$ amount.

| **Cost Tiers** | **ART ($)** | **Routine Care ($)** | | | **Labs ($)** |
| --- | --- | --- | --- | --- | --- |
|  |  | **CD4 >500/µL** | **CD4 200–500/µL** | **CD4 <200/µL** |  |
| **Low** | $20,900 | $1,700 | $2,300 | $17,100 | $300 |
| **Medium** | $35,000 | $1,700 | $2,300 | $17,100 | $300 |
| **High** | $45,000 | $1,700 | $2,300 | $17,100 | $300 |

Annual ART costs were inflated from their 2026 USD baseline values using Centers for Medicare & Medicaid Services (CMS) National Health Expenditure (NHE) projections: ART drug costs were inflated at 5.4% per year and routine care costs at 5.6% per year (CMS NHE 2026–2033 projections) [8].

Baseline ADAP spending was obtained from state-level reports in 2025 [9] and inflated to 2026 USD using the Medical Care Consumer Price Index. Because the ADAP budget has remained flat since 2014 [10], we assumed that the trend will continue to 2035 in the baseline scenario (ADAP continuation).

All projected costs and ADAP spending are discounted to the start of 2026 at 3% per year as per standard health economics convention [11].

**S1.3.	Primary outcome and jurisdictional correlates**

The primary outcome was the 2035 net cost-to-expenditure ratio (NCER), calculated as cumulative HIV care costs attributable to excess infections minus cumulative ADAP spending, divided by cumulative ADAP spending. Values greater than zero indicate that downstream care costs exceed avoided ADAP spending. We compared NCER by Medicaid expansion status and assessed Spearman correlations with the 2025 transmission rate, ADAP spending per client, the proportion of virally suppressed people with HIV receiving ADAP, and diagnosed HIV-weighted urbanicity. Jurisdictional inputs are reported in **Table S7** and the corresponding associations in **Figure 3** of the main manuscript.

#### S1.4. Simulation Procedure

To characterize uncertainty in epidemiologic and economic outcomes, we sampled 1,000 posterior JHEEM parameter sets representing uncertainty in model inputs, calibration targets, and fitted parameters. Each posterior model was evaluated under two policy scenarios: continuation of the baseline ADAP program and elimination of ADAP.

In the ADAP elimination scenario, we assumed that the absence of ADAP support would reduce the proportion of newly diagnosed individuals who initiate ART and achieve viral suppression. This reduction was modeled by sampling a viral suppression loss parameter ($L_{s,i})$from its corresponding probability distribution. The distribution was estimated from survey responses among clinicians and Ryan White program directors and was specified separately for Medicaid expansion and non-expansion states.

For the costing analysis, the proportion of newly diagnosed individuals who initiate ART and achieve viral suppression was taken directly from the ADAP elimination arm of the model rather than applied as a post hoc adjustment to a fixed baseline fraction. For each state $s$, posterior simulation $i$ (i = 1, …, 1000), and calendar year $t$ (t = 2026, …, 2035), the ART initiation fraction was calculated as

$$p_{s,i,t}^{\mathrm{ART} \mathrm{post}}=\frac{S_{s,i,t}}{D_{s,i,t}}$$

where $S_{s,i,t}$ is the modeled number of virally suppressed individuals and $D_{s,i,t}$ is the modeled diagnosed HIV prevalence under ADAP elimination, with the ratio bounded in [0,1]. Because both quantities are generated within the ADAP elimination scenario, this fraction already reflects the sampled loss of viral suppression $L_{s,i}$ and each state's reliance on ADAP, and no further adjustment was applied. The fraction therefore varies by state, simulation, and calendar year, allowing ART initiation among excess diagnoses to evolve over the projection period as the modeled care cascade responds to ADAP elimination. State-level values for this fraction at 2025 and 2035, under both ADAP continuation and elimination, are reported in **Table S5**.

Excess incident cases who did not initiate ART in the year of diagnosis were not assigned a subsequent probability of engagement, and delayed ART initiation was not modeled. These individuals accrue no HIV care costs at any point in the projection period, which yields a lower bound on downstream costs because it omits both the eventual treatment costs of those who would re-engage in care and the costs of untreated disease in the interim. Individuals who initiated ART were assumed to remain on treatment for the remainder of the horizon, so the treated excess population in each calendar year is the cumulative sum of ART initiations through that year.

The ADAP elimination scenario was simulated across 1,000 calibrated model runs, with loss of viral suppression sampled probabilistically. Each posterior draw was subsequently evaluated across three ART cost scenarios (low, medium, and high). This produced 3,000 economic evaluations across the 1,000 posterior draws. Results are reported as the median and 95% Uncertainty Ranges across all simulations. The uncertainty intervals reflect the combined effects of uncertainty in cost inputs, model parameters, and calibration targets.

#### S1.5. One-Way Threshold Analyses

To evaluate the robustness of the fiscal results to uncertainty in the epidemiologic consequences of ADAP elimination and the cost of ART, we conducted a complementary threshold analysis through 2035. The goal was to determine, for each state, how much viral suppression would need to be lost or how high ART cost would need to be for ADAP elimination to start costing more money than it saves.

First, we calculated the proportion of simulations that were cost-incurring in the primary analysis: For jurisdiction s, posterior simulation i, and ART-cost scenario k, we calculated the net cost-to-expenditure ratio as ${NCER}_{s,i,k}=\frac{C_{s,i,k}- A_{s}}{A_{s}}$, where is the discounted cumulative HIV care cost attributable to excess infections and is the discounted cumulative ADAP spending avoided through 2035. An NCER greater than zero indicates that downstream HIV care costs exceed the ADAP spending avoided, whereas an NCER below zero indicates net savings under the restricted healthcare-system perspective used in this analysis. For each jurisdiction, the probability that NCER exceeded zero was estimated as the proportion of economic evaluations satisfying. Under the primary specification, each jurisdiction contributed 3,000 economic evaluations, representing 1,000 posterior model simulations evaluated under each of the low, median, and high ART-cost scenarios. We pooled the three ART-cost scenarios with equal weight.

For the viral-suppression threshold analysis, we reran the JHEEM under fixed assumptions that 10%, 20%, …, 100% of ADAP-supported, virally suppressed clients would lose viral suppression following ADAP elimination beginning in 2026. For each jurisdiction, simulation, calendar year, and suppression-loss level, we calculated excess newly diagnosed cases relative to the corresponding no-intervention simulation. The proportion of newly diagnosed individuals initiating ART was calculated directly from the intervention-specific model outputs as the number of virally suppressed individuals divided by diagnosed HIV prevalence, bounded between zero and one. Because we did not model delayed ART initiation, annual ART initiations among excess cases equaled excess newly diagnosed cases multiplied by this intervention-specific ART-initiation fraction. Individuals initiating ART were assumed to remain on treatment, such that the number of excess individuals actively receiving ART in each year was the cumulative sum of ART initiations through that year.

For each suppression-loss level, the discounted cumulative HIV care cost was calculated as $C_{s,i,k}\left( l \right)= a_{k}D_{s,i}(l)+ R_{s,i}(l)$, where $l$ is the proportion losing viral suppression,$a_{k}$ is the 2026 annual ART drug cost under cost scenario, $D_{s,i}\left( l \right)$ is the cumulative number of discounted and $R_{s,i}(l)$ drug-cost-inflated excess ART person-years and is the corresponding discounted cumulative routine-care and laboratory cost. ART drug and routine-care costs were inflated annually by 5.4% and 5.6%, respectively, and discounted at 3% annually. At each of the ten simulated suppression-loss levels, we calculated the probability that NCER exceeded zero across all 3,000 economic evaluations. For visualization, we linearly interpolated the ten jurisdiction-specific probability estimates onto a density grid spanning 10%–100% suppression loss, producing the continuous heatmap shown in Panel B. We used this interpolation only to render the heatmap; it did not generate additional epidemiologic simulations or alter the probabilities estimated at the original grid points. We defined the jurisdiction-specific critical suppression-loss threshold as the level at which the probability of NCER exceeding zero reached 50%. When this threshold fell between two adjacent simulated suppression-loss levels, we estimated its location by linear interpolation.

For the ART-cost threshold analysis, we retained the complete draw-specific epidemiologic trajectory from each of the 1,000 posterior simulations under the primary ADAP-elimination specification. Each trajectory therefore retained its posterior epidemiologic parameter set and the randomly sampled, survey-informed viral-suppression loss value corresponding to the jurisdiction’s Medicaid expansion status. Epidemiologic outcomes consequently varied across posterior simulations but remained unchanged across the candidate ART-cost values evaluated within a given simulation. Within each trajectory, we varied the 2026 annual per-person ART drug cost over a logarithmic grid from $1000 to $100,000. Unlike Panel B, we calculated the probability that NCER exceeded zero directly at each ART-cost bin; we did not interpolate or statistically smooth these probabilities. For each jurisdiction and posterior simulation, cumulative attributable care cost at candidate annual drug cost $a$was calculated as $C_{s,i}= aD_{s,i}+ R_{s,i}$, where $D_{s,i}$represents discounted and drug-cost-inflated excess ART person-years and $R_{s,i}$ represents discounted cumulative routine-care and laboratory costs. The draw-specific break-even annual ART cost was therefore: $a_{s,i}^{*}=\frac{A_{s}-R_{s,i}}{D_{s,i}}$, because cumulative costs increase monotonically with the assumed annual ART cost, the jurisdiction-level cost at which the probability of NCER exceeding zero reached 50% was equivalent to the median draw-specific break-even cost across the 1,000 posterior simulations. If the 50% threshold fell outside the evaluated range, no critical threshold was reported

### S2. Projected State-Level HIV Epidemiology in 2025 and 2035, Pre- and Post- ADAP Elimination

We modeled ADAP elimination across 30 states and the District of Columbia. **Table S5** present projected HIV province, proportion of people with HIV virally suppressed, and number of people with HIV who are not virally suppressed at baseline (2025), and in 2035 under two scenarios: no ADAP elimination (continuation of status-que) and ADAP elimination in 2026.

**Table S5: Projected State-Level HIV Prevalence and Viral Suppression With and Without ADAP Elimination, 2025–2035.**

|  | **2025, Pre-ADAP elimination** | | | **2035, No ADAP Elimination** | | | **2035, Post-ADAP Elimination** | | |
| --- | --- | --- | --- | --- | --- | --- | --- | --- | --- |
| **State** | **HIV Prevalence** | **Prop of PWH Virally Suppressed** | **PWH living without Viral Suppression** | **HIV Prevalence** | **Prop of PWH Virally Suppressed** | **PWH living without Viral Suppression** | **HIV Prevalence** | **Prop of PWH Virally Suppressed** | **PWH living without Viral Suppression** |
| **AL** | 15,059 [14,575–15,363] | 74.6% [72.7–76.9%] | 3,824 [3,456–4,080] | 17,776 [16,049–18,852] | 82.6% [78.5–85.9%] | 3,059 [2,422–3,797] | 20,229 [17,503–22,213] | 61.1% [50.9–80.0%] | 7,876 [3,555–10,630] |
| **AR** | 7,058 [6,845–7,250] | 63.6% [61.1–66.1%] | 2,568 [2,380–2,768] | 9,572 [8,651–10,414] | 79.9% [74.4–83.4%] | 1,919 [1,550–2,493] | 10,097 [9,090–11,063] | 63.3% [48.1–78.6%] | 3,648 [2,048–5,502] |
| **AZ** | 20,151 [19,649–20,711] | 69.8% [66.4–73.3%] | 6,093 [5,444–6,780] | 23,841 [21,921–25,670] | 77.1% [68.3–83.5%] | 5,405 [3,920–7,606] | 24,973 [22,639–27,341] | 64.4% [49.6–78.7%] | 8,827 [5,153–13,085] |
| **CA** | 140,510 [137,494–143,542] | 76.8% [74.9–78.3%] | 32,649 [30,440–35,262] | 138,592 [132,291–146,072] | 82.9% [77.5–85.8%] | 23,609 [19,631–31,900] | 144,406 [135,410–154,286] | 68.0% [55.3–82.4%] | 46,221 [24,389–66,822] |
| **CO** | 15,168 [14,845–15,675] | 70.1% [67.6–72.4%] | 4,546 [4,183–4,945] | 19,735 [18,537–22,348] | 78.2% [71.9–82.7%] | 4,341 [3,349–5,601] | 21,620 [19,361–25,488] | 52.9% [30.7–77.0%] | 10,081 [4,615–16,605] |
| **DC** | 12,373 [12,062–12,686] | 62.5% [59.1–64.6%] | 4,634 [4,353–5,100] | 10,172 [9,472–10,886] | 68.5% [58.3–75.3%] | 3,182 [2,512–4,389] | 10,182 [9,475–10,897] | 65.5% [54.4–73.0%] | 3,494 [2,742–4,714] |
| **FL** | 126,300 [123,429–128,818] | 75.1% [73.4–76.7%] | 31,424 [29,341–33,511] | 138,721 [130,383–146,185] | 82.6% [79.2–85.2%] | 24,050 [20,077–29,011] | 147,186 [136,402–158,626] | 64.8% [57.3–80.3%] | 51,444 [26,965–65,421] |
| **GA** | 63,843 [62,494–65,290] | 75.2% [73.1–77.3%] | 15,841 [14,448–17,200] | 71,094 [66,168–76,613] | 81.2% [74.9–85.3%] | 13,442 [10,370–17,547] | 75,898 [69,817–81,931] | 65.4% [55.9–79.8%] | 26,339 [14,306–34,648] |
| **IL** | 35,671 [35,063–36,313] | 76.6% [74.0–78.3%] | 8,352 [7,692–9,279] | 33,638 [31,876–35,125] | 84.8% [81.1–87.3%] | 5,122 [4,319–6,353] | 36,402 [33,113–40,097] | 64.8% [47.2–83.3%] | 12,921 [5,670–20,405] |
| **IN** | 12,737 [12,416–13,187] | 72.9% [70.9–74.5%] | 3,452 [3,230–3,784] | 14,061 [12,664–15,650] | 80.5% [75.2–84.2%] | 2,730 [2,177–3,595] | 14,572 [12,933–16,401] | 63.4% [46.7–79.6%] | 5,384 [2,798–8,143] |
| **KY** | 8,821 [8,620–9,050] | 75.6% [73.2–77.5%] | 2,155 [1,985–2,376] | 9,681 [9,126–10,433] | 86.7% [84.2–88.4%] | 1,291 [1,111–1,557] | 10,603 [9,527–12,093] | 61.7% [39.2–84.5%] | 4,033 [1,521–7,075] |
| **LA** | 22,324 [21,657–22,797] | 80.3% [78.3–81.5%] | 4,410 [4,062–4,856] | 24,403 [22,531–26,197] | 86.9% [83.6–88.3%] | 3,212 [2,729–3,955] | 25,621 [23,400–27,798] | 73.6% [62.0–85.9%] | 6,795 [3,473–10,178] |
| **MA** | 20,987 [20,424–21,503] | 72.3% [69.5–74.4%] | 5,797 [5,318–6,469] | 21,403 [19,660–22,753] | 75.0% [66.4–80.6%] | 5,342 [3,934–7,235] | 22,087 [20,100–23,998] | 55.6% [36.3–74.7%] | 9,808 [5,357–14,732] |
| **MD** | 33,130 [31,806–36,579] | 79.3% [76.6–80.6%] | 7,133 [6,259–7,680] | 31,579 [27,817–38,818] | 87.6% [83.6–88.6%] | 4,223 [3,270–4,874] | 32,244 [28,237–39,759] | 75.5% [65.0–86.6%] | 7,960 [4,227–12,281] |
| **MI** | 17,613 [17,178–18,006] | 84.5% [83.2–85.3%] | 2,729 [2,573–2,998] | 17,596 [16,359–18,907] | 88.9% [87.0–89.4%] | 1,960 [1,773–2,389] | 18,112 [16,796–19,567] | 75.2% [64.2–87.5%] | 4,465 [2,176–6,718] |
| **MN** | 9,975 [9,756–10,214] | 80.2% [78.3–82.5%] | 1,973 [1,747–2,166] | 12,300 [11,571–13,127] | 82.7% [78.7–85.7%] | 2,137 [1,738–2,652] | 13,186 [11,981–14,598] | 69.6% [57.2–82.2%] | 4,039 [2,203–6,048] |
| **MO** | 13,815 [13,462–14,158] | 76.0% [73.7–78.6%] | 3,318 [2,946–3,662] | 15,372 [13,941–16,755] | 78.2% [73.4–83.5%] | 3,317 [2,568–4,160] | 19,247 [15,309–23,500] | 59.0% [41.1–77.5%] | 7,772 [3,545–13,139] |
| **MS** | 10,160 [9,897–10,380] | 75.7% [74.0–77.4%] | 2,470 [2,283–2,638] | 11,382 [10,662–12,119] | 85.2% [82.3–87.1%] | 1,689 [1,426–2,065] | 12,067 [11,207–13,009] | 64.9% [57.3–82.4%] | 4,256 [2,026–5,389] |
| **NC** | 37,398 [36,479–38,220] | 73.8% [72.0–75.9%] | 9,769 [8,977–10,511] | 44,747 [41,291–48,042] | 81.3% [76.9–84.6%] | 8,319 [6,775–10,558] | 47,968 [43,142–52,484] | 63.3% [46.9–80.6%] | 17,705 [8,791–26,998] |
| **NJ** | 35,840 [35,325–36,294] | 59.3% [55.4–63.5%] | 14,606 [13,061–15,941] | 35,921 [33,923–37,785] | 67.8% [52.5–78.5%] | 11,552 [7,680–17,252] | 36,630 [34,468–38,836] | 59.3% [42.0–73.6%] | 14,878 [9,412–21,870] |
| **NV** | 13,260 [12,917–13,706] | 68.2% [64.5–70.8%] | 4,244 [3,819–4,722] | 18,035 [16,742–19,844] | 78.2% [66.1–82.5%] | 4,074 [3,030–6,171] | 18,848 [17,236–20,661] | 61.8% [45.0–78.1%] | 7,211 [3,879–10,677] |
| **NY** | 122,563 [120,304–125,403] | 72.7% [70.6–74.7%] | 33,505 [30,985–35,941] | 118,503 [112,898–125,274] | 77.5% [71.0–82.1%] | 26,806 [21,256–33,979] | 121,642 [114,640–129,349] | 67.6% [57.4–78.4%] | 39,408 [25,708–53,306] |
| **OH** | 26,064 [25,477–26,671] | 76.3% [74.6–77.9%] | 6,190 [5,764–6,650] | 29,697 [27,885–31,888] | 86.2% [83.9–87.8%] | 4,101 [3,564–4,843] | 31,207 [28,526–34,295] | 73.1% [61.6–85.0%] | 8,383 [4,477–12,580] |
| **OK** | 7,303 [7,152–7,466] | 65.8% [63.3–68.7%] | 2,503 [2,288–2,666] | 9,215 [8,391–10,029] | 72.0% [64.6–77.9%] | 2,586 [2,030–3,214] | 9,705 [8,716–10,896] | 52.6% [34.1–72.3%] | 4,595 [2,611–6,690] |
| **PA** | 38,226 [37,360–39,015] | 68.8% [66.7–71.0%] | 11,941 [11,032–12,755] | 39,816 [37,769–42,167] | 79.0% [72.3–84.6%] | 8,419 [6,078–11,000] | 40,773 [38,365–43,644] | 67.9% [55.7–79.9%] | 13,186 [8,019–18,489] |
| **SC** | 18,578 [18,158–18,931] | 77.8% [76.7–79.3%] | 4,137 [3,867–4,323] | 20,480 [19,598–22,188] | 85.1% [83.0–86.6%] | 3,080 [2,738–3,542] | 22,458 [20,629–24,284] | 59.9% [50.4–81.2%] | 9,040 [3,870–11,591] |
| **TN** | 19,783 [19,392–20,209] | 67.4% [65.3–69.4%] | 6,462 [5,995–6,901] | 23,877 [21,974–25,941] | 77.6% [71.0–83.5%] | 5,394 [3,716–7,347] | 27,753 [23,818–31,266] | 42.0% [26.7–72.1%] | 16,224 [6,703–22,202] |
| **TX** | 107,123 [104,995–109,781] | 71.1% [68.7–73.5%] | 31,065 [28,282–33,671] | 121,408 [113,909–130,925] | 74.3% [65.5–81.2%] | 31,198 [22,399–41,788] | 129,917 [119,127–143,486] | 63.6% [51.8–75.8%] | 47,288 [29,795–64,505] |
| **VA** | 25,511 [24,939–26,089] | 79.0% [77.3–80.2%] | 5,362 [5,045–5,799] | 27,930 [26,064–30,019] | 87.9% [86.4–88.8%] | 3,376 [3,092–3,842] | 29,520 [26,891–32,582] | 72.2% [58.9–86.3%] | 8,170 [3,783–12,995] |
| **WA** | 16,144 [15,744–16,580] | 86.0% [84.9–86.8%] | 2,262 [2,127–2,445] | 21,175 [19,527–22,652] | 88.7% [87.2–89.3%] | 2,397 [2,172–2,761] | 22,298 [20,270–24,622] | 70.2% [54.2–86.7%] | 6,769 [2,775–10,873] |
| **WI** | 7,256 [7,095–7,447] | 84.2% [83.0–85.3%] | 1,144 [1,064–1,227] | 8,898 [8,072–9,632] | 88.6% [87.4–89.2%] | 1,023 [913–1,130] | 9,953 [8,866–10,960] | 66.6% [58.8–85.3%] | 3,343 [1,323–4,402] |
| **Total (US)** | **1,061,315 [1,054,695–1,068,391]** | **73.9%**  **[73.4–74.5%]** | **276,541 [270,795–282,220]** | **1,142,838 [1,122,115–1,160,405]** | **80.3%**  **[78.9–81.7%]** | **224,986**  **[208,240–241,140]** | **1,210,440 [1,145,573–1,260,504]** | **65.0%**  **[55.4–78.9%]** | **425,163 [243,220–554,711]** |

### S3. Projected Annual Outcomes and Costs Across the United States

We modeled ADAP elimination across 30 states and the District of Columbia. Presenting the US aggregate epidemiologic and economic outcomes side by side provides a concrete benchmark for interpreting the model results and demonstrates the potential public health and cost consequences of ADAP reductions (**Table S6**). Complete results for all modeled jurisdictions are reported in **Table 1** in the main manuscript.

**Table S6. Projected Excess HIV Diagnoses, ART Usage, and Costs of Care Following an ADAP Elimination Across the United States, 2026–2035.** The table summarizes projected new diagnoses and ART usage resulting from excess HIV infections after ADAP elimination and resulting HIV care costs from a healthcare perspective, compared to baseline (ADAP continuation). Care costs include routine care, laboratory monitoring, and ART expenditures, reported annually and cumulatively. ADAP spending in the baseline scenario is reported in parallel to estimated costs. The net cost is calculated as cumulative HIV care costs minus cumulative ADAP spending. Finally, the net cost to expenditure ratio (NCER) is calculated as net cost over ADAP spending. Values represent median [95% uncertainty range] across all simulations.

| **Year** | **Excess New Infections** | **New Diagnoses** | **Total Number Initiating ART** | **Person-Years on ART** | **Annual HIV Care Cost for Excess Infections** | **Cumulative HIV Care Cost for Excess Infections** | **Annual ADAP Spending** | **Cumulative ADAP Spending** | **Net Cost (Cum HIV Care Cost for Excess Infections − Cum ADAP Spending)** | **NCER** |
| --- | --- | --- | --- | --- | --- | --- | --- | --- | --- | --- |
| **2026** | 6,243 [735–9,767] | 1,311 [155–2,102] | 846 [114–1,237] | 846 [114–1,237] | $27.7M [$3.8M–$57.9M] | $27.7M [$3.8M–$57.9M] | $748.5M | $748.5M | −$720.8M [−$744.7M–−$690.6M] | −0.963 [−0.995–−0.923] |
| **2027** | 18,048 [2,131–28,213] | 6,930 [822–11,005] | 3,426 [495–4,727] | 5,127 [723–7,198] | $140.3M [$20.9M–$285.9M] | $168.1M [$24.7M–$343.9M] | $748.5M | $1.48B | −$1.31B [−$1.45B–−$1.13B] | −0.886 [−0.983–−0.767] |
| **2028** | 31,427 [3,718–49,168] | 15,429 [1,869–24,533] | 5,305 [776–7,240] | 14,685 [2,103–20,418] | $320.8M [$48.0M–$652.8M] | $488.6M [$72.8M–$995.2M] | $748.5M | $2.18B | −$1.69B [−$2.11B–−$1.19B] | −0.776 [−0.967–−0.544] |
| **2029** | 45,903 [5,425–71,762] | 25,922 [3,149–41,027] | 6,525 [946–8,895] | 30,745 [4,462–42,491] | $552.6M [$82.5M–$1.12B] | $1.04B [$155.2M–$2.12B] | $748.5M | $2.87B | −$1.82B [−$2.71B–−$749.4M] | −0.637 [−0.946–−0.261] |
| **2030** | 61,041 [7,216–95,747] | 37,618 [4,546–59,471] | 7,395 [1,062–10,087] | 54,210 [7,922–74,553] | $826.4M [$123.0M–$1.67B] | $1.87B [$278.2M–$3.79B] | $748.5M | $3.53B | −$1.66B [−$3.25B–$254.8M] | −0.471 [−0.921–0.072] |
| **2031** | 76,822 [9,072–120,608] | 50,252 [6,045–79,404] | 8,044 [1,152–10,993] | 85,696 [12,484–117,564] | $1.14B [$168.6M–$2.29B] | $3.00B [$447.0M–$6.07B] | $748.5M | $4.18B | −$1.17B [−$3.73B–$1.89B] | −0.281 [−0.893–0.454] |
| **2032** | 93,006 [10,975–146,245] | 63,505 [7,616–100,557] | 8,524 [1,221–11,676] | 125,654 [18,258–172,224] | $1.48B [$219.0M–$2.97B] | $4.48B [$665.8M–$9.03B] | $748.5M | $4.80B | −$324.6M [−$4.14B–$4.23B] | −0.068 [−0.861–0.881] |
| **2033** | 109,454 [12,913–172,441] | 77,103 [9,243–122,434] | 8,891 [1,280–12,235] | 174,727 [25,294–239,130] | $1.85B [$273.8M–$3.72B] | $6.33B [$939.6M–$12.75B] | $748.5M | $5.41B | $919.8M [−$4.47B–$7.34B] | 0.170 [−0.826–1.356] |
| **2034** | 126,242 [14,914–199,413] | 91,338 [10,899–144,885] | 9,189 [1,323–12,689] | 232,809 [33,624–318,781] | $2.24B [$332.1M–$4.53B] | $8.57B [$1.27B–$17.27B] | $748.5M | $6.00B | $2.57B [−$4.73B–$11.26B] | 0.428 [−0.788–1.876] |
| **2035** | 143,347 [16,946–226,886] | 105,730 [12,599–167,856] | 9,436 [1,358–13,099] | 300,551 [43,277–411,370] | $2.67B [$393.7M–$5.39B] | $11.24B [$1.67B–$22.66B] | $748.5M | $6.58B | $4.66B [−$4.91B–$16.09B] | 0.709 [−0.746–2.446] |

### S4. Projected Outcomes and Cost Summary Across all Jurisdictions

Complete results for all modeled jurisdictions are reported in **Table 1** of the main manuscript. **Table S7** provides contextual characteristics for the modeled jurisdictions, including Medicaid expansion status, total ADAP expenditures and expenditures per client, the proportion of people with HIV who are virally suppressed and receiving ADAP, the average HIV transmission rate (calibrated), and the number of diagnosed people with HIV living in urban areas (the urbanicity metric). These characteristics facilitate interpretation of the heterogeneity in projected epidemiologic and economic outcomes across jurisdictions.

**Table S7. Characteristics of Modeled Jurisdictions included in the analysis.** Values represent the median values across 3,000 simulated economic evaluations in each state. This table presents the state-level values underlying Figure 2: NCER at 2035 and its associated state characteristics (proportion of suppressed people with HIV on ADAP, average transmission rate, viral suppression, and diagnosed HIV-weighted urbanicity), by Medicaid expansion status.

| **State** | **Medicaid Expansion Status** | **NCER** | **ADAP Spending per Client (2026 USD)** | **Cum. ADAP Spending through 2035 (2026 USD)** | **Proportion of Suppressed PWH on ADAP (%)** | **Average Transmission Rate** | **Diagnosed HIV-weighted Urbanicity** |
| --- | --- | --- | --- | --- | --- | --- | --- |
| **Missouri** | Medicaid expansion | 3.792 | $1,980 | $88,911,408 | 44.1 | 1.030 | 0.858 |
| **Alabama** | Non-expansion | 3.100 | $2,389 | $95,321,538 | 33.0 | 0.689 | 0.682 |
| **Tennessee** | Non-expansion | 2.912 | $1,487 | $124,440,743 | 61.3 | 1.591 | 0.821 |
| **Wisconsin** | Non-expansion | 2.370 | $2,171 | $42,090,433 | 33.1 | 2.273 | 0.833 |
| **South Carolina** | Non-expansion | 1.772 | $2,091 | $119,156,637 | 40.0 | 1.018 | 0.684 |
| **Mississippi** | Non-expansion | 1.395 | $2,595 | $68,299,190 | 30.9 | 0.744 | 0.543 |
| **Florida** | Non-expansion | 1.335 | $2,810 | $751,897,125 | 29.5 | 1.041 | 0.943 |
| **Kentucky** | Medicaid expansion | 1.249 | $1,170 | $51,287,301 | 59.0 | 1.259 | 0.816 |
| **Colorado** | Medicaid expansion | 1.244 | $1,372 | $83,878,928 | 49.5 | 0.919 | 0.923 |
| **North Carolina** | Non-expansion | 1.220 | $1,870 | $210,728,972 | 41.4 | 1.071 | 0.751 |
| **Minnesota** | Medicaid expansion | 1.194 | $2,206 | $56,911,101 | 29.1 | 3.484 | 0.885 |
| **Arkansas** | Medicaid expansion | 1.171 | $2,142 | $41,272,661 | 40.5 | 0.425 | 0.651 |
| **Washington** | Medicaid expansion | 1.070 | $1,754 | $88,565,429 | 38.0 | 3.161 | 0.888 |
| **Georgia** | Non-expansion | 1.044 | $2,743 | $361,961,401 | 26.3 | 0.834 | 0.866 |
| **Texas** | Non-expansion | 1.033 | $2,748 | $662,954,266 | 26.7 | 1.107 | 0.904 |
| **Louisiana** | Medicaid expansion | 1.010 | $3,074 | $148,118,579 | 28.0 | 0.573 | 0.800 |
| **Virginia** | Medicaid expansion | 0.931 | $2,315 | $160,026,694 | 34.1 | 2.168 | 0.857 |
| **Illinois** | Medicaid expansion | 0.898 | $2,092 | $249,755,883 | 45.8 | 2.425 | 0.958 |
| **Ohio** | Medicaid expansion | 0.597 | $2,676 | $152,408,771 | 30.4 | 1.355 | 0.887 |
| **Nevada** | Medicaid expansion | 0.588 | $2,053 | $66,850,257 | 36.9 | 0.930 | 0.974 |
| **Arizona** | Medicaid expansion | 0.490 | $2,713 | $112,557,793 | 30.7 | 2.018 | 0.920 |
| **Oklahoma** | Medicaid expansion | 0.489 | $1,979 | $44,889,755 | 40.2 | 1.233 | 0.771 |
| **Indiana** | Medicaid expansion | 0.295 | $2,126 | $79,997,626 | 39.9 | 1.278 | 0.846 |
| **Michigan** | Medicaid expansion | 0.287 | $3,071 | $116,193,764 | 26.6 | 2.430 | 0.858 |
| **Massachusetts** | Medicaid expansion | -0.017 | $1,855 | $130,391,653 | 47.4 | 1.794 | 0.934 |
| **California** | Medicaid expansion | -0.079 | $2,729 | $917,050,671 | 32.0 | 0.791 | 0.964 |
| **Maryland** | Medicaid expansion | -0.107 | $3,165 | $207,891,912 | 24.3 | 0.904 | 0.933 |
| **Pennsylvania** | Medicaid expansion | -0.199 | $2,946 | $225,738,355 | 26.9 | 0.597 | 0.892 |
| **New York** | Medicaid expansion | -0.227 | $3,887 | $787,533,423 | 23.7 | 1.161 | 0.960 |
| **New Jersey** | Medicaid expansion | -0.268 | $3,992 | $236,822,403 | 24.1 | 0.785 | 0.962 |
| **District of Columbia** | Medicaid expansion | -0.967 | $8,866 | $92,710,931 | 3.3 | 0.407 | 1.000 |

### S5. Projected State-Level Outcomes and Contextual Characteristics

**Figure S2** presents correlations between the primary outcome, the net cost-to-expenditure ratio (NCER) through 2035, and four jurisdiction-level characteristics: projected cumulative ADAP expenditures through 2035 (reported in 2026 USD), viral suppression among diagnosed people with HIV, diagnosed HIV prevalence per capita, and the proportion of ADAP recipients who are virally suppressed. Point size represents the number of ADAP clients at the 2025 baseline in panel A and cumulative ADAP spending through 2035 in panels B–D. Each panel illustrates the direction and strength of the association between NCER and the corresponding characteristic across the modeled jurisdictions.

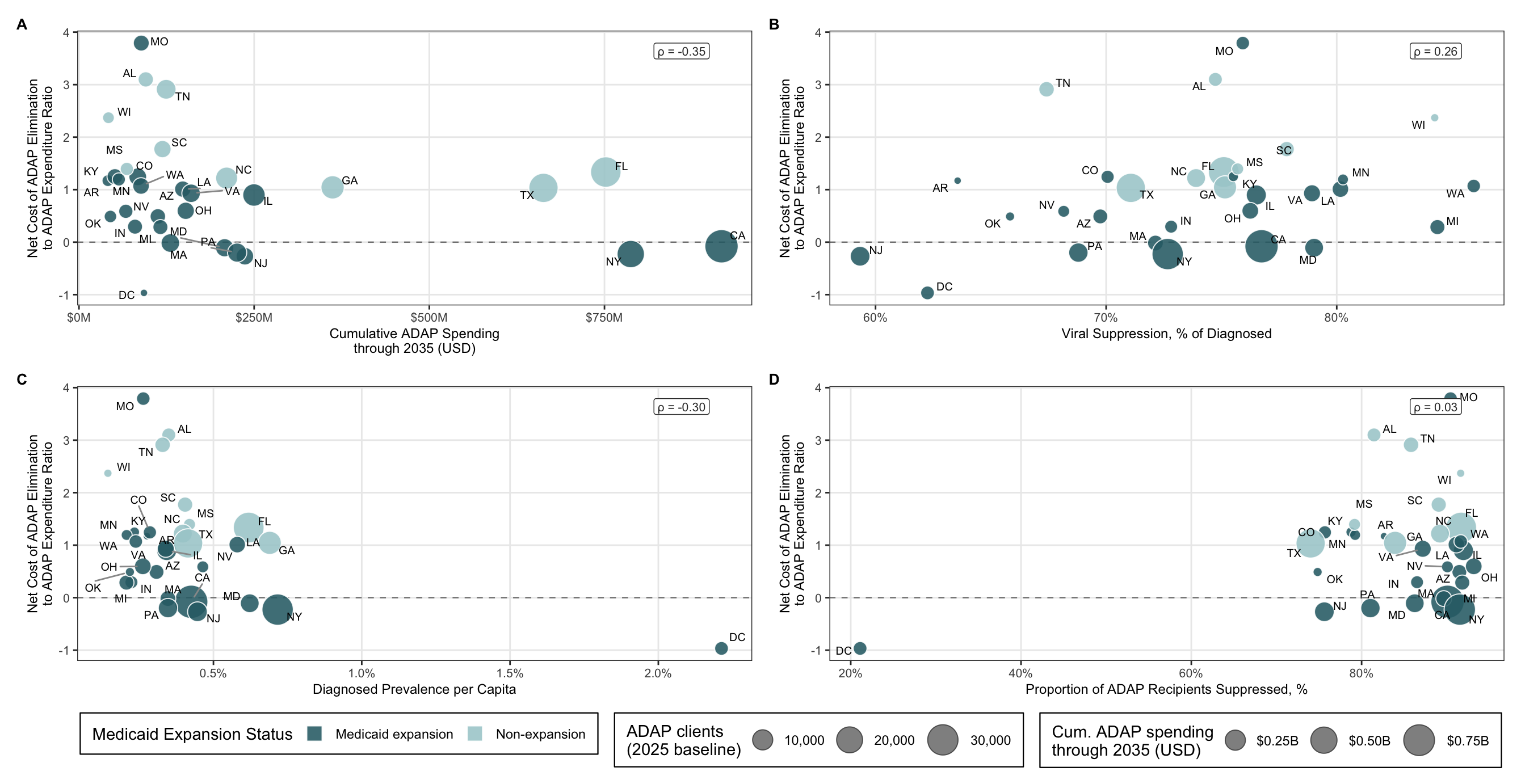

**Figure S2. Associations between the projected net cost-to-expenditure ratio (NCER) of ADAP elimination and jurisdiction-level characteristics across 30 US states and the District of Columbia.** Panels show NCER through 2035 in relation to (**A**) cumulative ADAP spending through 2035, (**B**) viral suppression among diagnosed people with HIV, (**C**) diagnosed HIV prevalence per capita, and (**D**) the proportion of ADAP recipients who are virally suppressed. Each point represents one jurisdiction, labeled by its abbreviation. **In panel A, point size is proportional to the number of ADAP clients at the 2025 baseline. In panels B–D, point size is proportional to cumulative ADAP spending through 2035.** Point color indicates Medicaid expansion status. The symbol ρ denotes Spearman’s rank correlation coefficient. The dashed horizontal line marks cost-neutrality (NCER = 0); values above zero indicate that projected downstream HIV care costs exceed ADAP spending averted by elimination.

### S6. Threshold Analyses Results

Under the primary specification, NCER exceeded zero in at least half of economic evaluations in 24 of 31 jurisdictions and in at least 80% of evaluations in 17 jurisdictions (**Figure S4A**). The probability was highest in Tennessee (98%), followed by Alabama and Wisconsin (96% each), Missouri (95%), and South Carolina (94%). The District of Columbia was the only jurisdiction with NCER below zero across all evaluations.

Across Medicaid non-expansion states, the critical viral-suppression-loss threshold ranged from 13% to 29%, well below the survey-informed median loss of 77.6% (**Figure S4B**). Thresholds were more heterogeneous among Medicaid expansion jurisdictions (17%-71%). Tennessee and Missouri remained cost-incurring across all evaluated suppression-loss levels, whereas the District of Columbia remained cost-saving throughout.

Critical annual ART-cost thresholds ranged from $4,600 to $12,800 across non-expansion states, below the primary low-cost scenario of $20,900 in all eight jurisdictions (**Figure S4C**). Among expansion jurisdictions, thresholds ranged from $3,300 to $43,500. These threshold analyses indicate that the fiscal findings were not driven solely by extreme model draws or a single assumption about viral-suppression loss or ART cost.

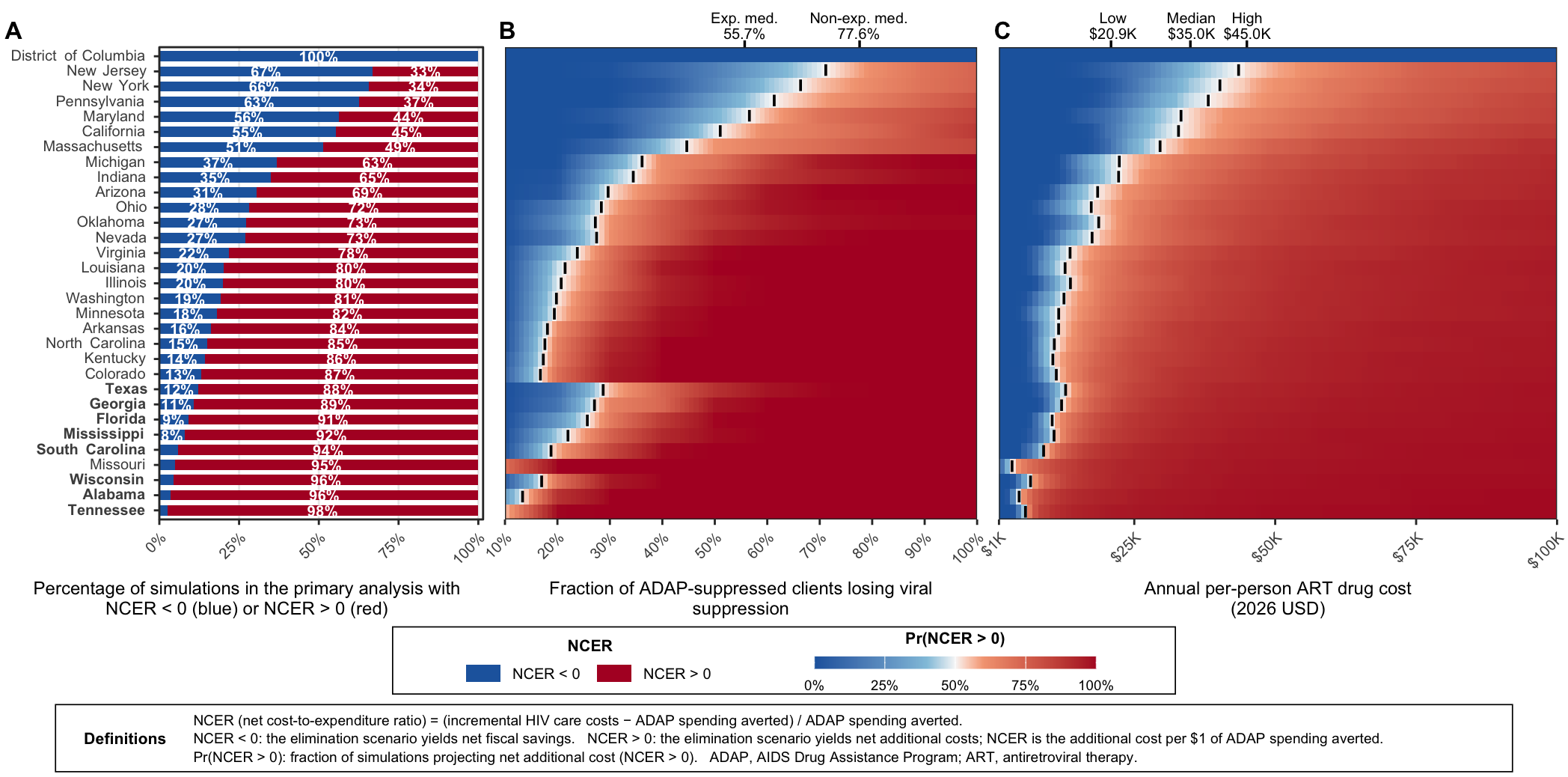

**Figure S3. One-way threshold analysis of viral suppression and ART costs on NCER through 2035.** NCER represents the projected (HIV care costs − savings) / savings under the ADAP elimination scenario, where savings reflect ADAP spending averted by elimination. NCER > 0 (red) indicates a cost-incurring outcome, in which the health system incurs additional cost per dollar of ADAP spending averted; NCER < 0 (blue) indicates a cost-saving outcome, in which averted ADAP spending exceeds the resulting increase in HIV care costs by 2035. (A) State-specific probabilistic results under the primary specification, calculated across all 1,000 posterior simulations. Jurisdictions are grouped by Medicaid expansion status and ordered within each group by the probability that NCER exceeds zero. (B) Threshold behavior across a fixed range of ADAP-attributable viral suppression losses (10%-100%). The upper axis identifies the medians of the fitted survey distributions for Medicaid expansion states (55.7%) and non-expansion states (77.6%). (C) Threshold behavior across a range of annual per-person ART drug costs, ranging from $1000 to $100,000. The upper axis marks the low ($20,900), median ($35,000), and high ($45,000) ART cost levels used in the primary analysis. In panels B and C, color represents the probability that NCER exceeds zero, with white centered at 50%. Black tick marks within each jurisdictional row identify the interpolated grid value at which this probability reaches 50%.

7. Office of Procurement. Acquisition and logistics (opal). Federal supply schedule. U.S. Department of Veterans Affairs. https://www.va.gov/opal/nac/fss/pharmprices.asp

8. Centers for Medicare & Medicaid Services, Office of the Actuary. National Health Expenditure Projections 2023–2033. Baltimore, MD: CMS; 2024.

9. Health Resources & Services Administration. FY25 Ryan White HIV/AIDS Program Part B Grant Awards. U.S. Department of Health & Human Services; January 2026. https://ryanwhite.hrsa.gov/about/parts-and-initiatives/part-b-adap/fy-2025-grant-awards

10. Dawson L, Kates J. Constrained budgets lead states to restrict HIV drug access through Ryan White. KFF. March 2, 2026. https://www.kff.org/hiv-aids/constrained-budgets-lead-states-to-restrict-hiv-drug-access-through-ryan-white/

11. Gold MR, Siegel JE, Russell LB, Weinstein MC, eds. Cost-Effectiveness in Health and Medicine. New York: Oxford University Press; 1996.

.
